# Peripheral artery disease affects the legs of claudicating patients in a diffuse manner irrespective of the level of the arterial tree primarily involved

**DOI:** 10.1101/2022.02.14.22270963

**Authors:** Todd J. Leutzinger, Panagiotis Koutakis, Matthew A. Fuglestad, Hafizur Rahman, Holly Despiegelaere, Mahdi Hassan, Molly Schieber, Jason M. Johanning, Nick Stergiou, G. Matthew Longo, George P. Casale, Sara A. Myers, Iraklis I. Pipinos

## Abstract

**Objective:** Different levels of arterial occlusive disease (aortoiliac, femoropopliteal, multi-level disease) can produce claudication symptoms in different leg muscle groups (buttocks, thighs, calves) in patients with peripheral artery disease (PAD). We tested the hypothesis that different levels of occlusive disease uniquely affect the muscles of PAD legs and produce distinctive patterns in the way claudicating patients walk.

**Methods:** 97 PAD patients and 35 healthy controls were recruited. PAD patients were categorized to aortoiliac, femoropopliteal and multi-level disease groups using computerized tomographic angiography. Subjects performed walking trials both pain-free and during claudication and joint kinematics, kinetics, and spatiotemporal parameters were calculated to evaluate the net contribution of the calf, thigh and buttock muscles.

**Results:** PAD patients with different levels of arterial occlusions had different patterns of symptoms in their calves, thighs and buttocks. However, no significant biomechanical differences were found between PAD groups during the pain-free conditions with minimal differences between PAD groups in the claudicating state. All statistical differences in the pain-free condition occurred between healthy controls and one or more PAD groups. A discriminant analysis function was able to adequately predict if a subject was a control with over 70% accuracy, but the function was unable to differentiate between PAD groups.

**Conclusions:** In-depth gait analyses of claudicating PAD patients indicate that different levels of arterial disease produce symptoms that affect different muscle groups across the lower extremity but impact the function of the leg muscles in a diffuse manner generating similar walking impairments.

## INTRODUCTION

Intermittent claudication is the most common manifestation of peripheral artery disease (PAD)[1]. Claudication is defined as walking-induced, ischemic leg pain that is most commonly described by the patient as severe muscle cramping, tightness, aching, and/or fatigue[2]. Claudication results in severely decreased functional ability[3] and quality of life[4] and is associated with increased risk for the occurrence of other cardiovascular related events and mortality[5]–[7].

Patients with PAD walk with an abnormal gait pattern compared to healthy, age matched controls[8], [9]. The majority of these gait abnormalities are present from the first step the patients take, well before the onset of claudication, and become more severe when claudication discomfort begins[10], [11]. Research from our group and others has shown that the abnormal gait pattern is characterized by slower walking speed, decreased cadence, increased stance time, and shorter stride lengths compared to healthy, age-matched controls[12]. Furthermore, patients with PAD walk with altered joint angles[12], [13] and reduced joint torques[10], [14] and powers[10], [14], [15] around the ankle, knee, and hip joints compared to their healthy counterparts.

Atherosclerotic stenoses/occlusions can occur at any or multiple levels throughout the arterial tree supplying the leg, restricting blood flow to the muscles responsible for movement at the hip, knee, and ankle joints. These blockages are typically grouped into the following general levels: aortoiliac (AI), femoropopliteal (FP), tibial, and multi-level disease (MLD)[16]. Aortoiliac disease can produce claudication symptoms in all major muscle groups (calves, thighs and buttocks) of the leg [17], [18]. Femoropopliteal disease can produce claudication symptoms in the muscles of the calves[17]. Tibial disease may also produce claudication symptoms in the muscles of the calf. Multi-level atherosclerosis is associated with vascular narrowing/occlusion in more than one level of the arterial tree of the leg. Patients with MLD tend to have worse hemodynamic compromise, than patients with single level disease, and they can experience claudication symptoms in all major muscle groups of the leg including those of the calves, thighs and buttocks. However, they usually experience claudication pain distal to the most affected arterial segment[18] [19].

Arterial blockages and the ischemia and ischemia/reperfusion they generate in the muscles they affect, produce an ischemic myopathy in the legs of PAD patients. At the histological level the principal characteristics of the myopathy are myofiber degeneration and fibrosis of the extracellular matrix and microvessels of the affected muscles[20]–[25], [64]. At the biochemical level the myopathy is characterized by oxidative damage, mitochondrial dysfunction, cytoskeletal disintegration and a characteristic upregulation of cytokines[20], [22], [26]–[31]. The myopathy of PAD in association with exercise-induced ischemia produce the, familiar to all vascular practitioners, abnormal muscle performance and metabolically inefficient walking pattern of claudicating patients[10]–[12], [14], [15], [32]. Because the level of occlusive disease usually determines the muscle group that is affected by the discomfort of claudication, it is possible that disease level may also play a primary role in the patient’s walking biomechanics and determine a patient’s compensation strategies during gait. Specifically, it is likely that different levels of occlusive disease produce ischemia, myopathy, and functional compromise of different muscle groups in the PAD legs which then results in distinctive gait patterns. It is also possible that as different muscle groups are affected, the patient’s muscular and neurological systems respond by alleviating the load placed on the more affected muscle groups while increasing the contribution of less affected muscles in an attempt to reduce pain and improve metabolic efficiency. Investigating the influence of disease level on a patient’s gait may provide pivotal information regarding the pathophysiology of claudication and the development of each patient’s walking impairments. By identifying if and how the level of vascular stenosis/occlusion affects the contributions of the muscles that move particular joints, and results in specific gait abnormalities in individuals with PAD, clinicians and therapists may be better able to understand the type and degree of functional limitation produced by different patterns of occlusive disease. This knowledge may help with the development of treatment plans that target specific muscles such as the ankle or knee or hip flexors/extensors based on the vascular stenosis/occlusion level and may provide the basis for physical therapy and exercise interventions that are more patient-specific and possibly more effective in rehabilitating each patient’s PAD limitations.

In this study we tested the hypothesis that the level of arterial stenosis/occlusion would uniquely impair the net muscle contributions, walking pattern and overall walking ability between patients with different levels of disease and compared to healthy controls. More specifically we aimed to determine if different levels of vascular stenosis/ occlusion uniquely affect walking patterns, as described by objective kinematic, kinetic, spatiotemporal gait parameters, and six minute walk distances and by subjective measurements acquired through the Walking Impairment Questionnaire (WIQ) and the Medical Outcomes Study Short Form 36 Health Survey (SF-36). Furthermore, we sought to determine if claudicating patients can be classified according to their level of disease based on their pattern of gait impairments.

## METHODS

### Study Participants

Ninety Seven patients (96 males, 1 female) diagnosed with claudication secondary to PAD, age (63.34 ± 6.49 years), height (1.76 ± 0.07 m), and body-mass (87.31± 17.22 kg) were recruited from the Nebraska and Western Iowa Veterans Affairs Medical Center and the University of Nebraska Medical Center. Thirty-five healthy controls with similar age (65.23 ± 9.51 years), height (1.74 ± 0.08 m), and body-mass (79.90± 13.80 kg) were recruited from the community.

### Inclusion and Exclusion Criteria

Patient inclusion criteria included the ability to provide written, informed consent for study participation, a history of chronic, exercise-limiting claudication, an ABI ≤ 0.90 at rest[33], [34] and stable blood pressure, lipid, and/or diabetes regimens and risk factor control for six weeks or more. Patient exclusion criteria included ischemic rest pain, tissue loss due to PAD, the presence of acute lower leg ischemia and significant limitations in the ability to walk due to conditions other than claudication such as pathology occurring in the leg (joint, musculoskeletal, neurological), heart, or lungs. Patients were screened and evaluated by one of two board certified vascular surgeons prior to study enrollment. Healthy control participants were excluded for the same exclusion criteria as the PAD population or if they had walking limitations, if they experienced leg pain during walking, or if their ABI values at rest were ≤ 0.90. All controls were screened by a trained member of the study team measuring systolic pressures at the brachial and pedal arteries using Doppler [6]. Informed consent was obtained from all study participants prior to data collection according to the guidelines of the institutional review boards of the two medical centers.

### CTA and determination of level of disease

Patients with PAD underwent computerized tomographic angiography (CTA) of the abdomen, pelvis and lower extremities including delayed imaging. 1.5-mm cross-sectional images were obtained from the lower chest to the toes and were used by one of two board certified vascular surgeons to determine the level of disease[35] [36]. The degree of arterial stenosis was considered minimal if the arterial diameter was decreased by less than 10%, mild if the arterial diameter was decreased by 10 to 30%, moderate if the arterial diameter was decreased by 30% to 70%, and severe if the arterial diameter was decreased by more than 70%. A patient was considered as having disease in a segment of his arterial tree if there was evidence of moderate or severe stenosis (30-100% blockage) in one or more arteries of that segment. Patients with minimal (<10% diameter reduction) or mild stenosis (10-30% diameter reduction) in the arteries of a particular segment were considered as not having occlusive disease in that segment. Occlusive disease patterns were described as Aortoiliac (AI, stenosis/occlusion in the aorta, iliac and common femoral arteries), Femoropopliteal (FP, stenosis/occlusion in the superficial femoral and popliteal arteries) and multilevel disease (MLD, Aortoiliac + femoropopliteal disease). A total of 26 (of 97) PAD patients had significant disease that affected one to three of their tibial arteries but all of them had dominant disease in the more proximal AI or FP segments and were analyzed as part of the dominant proximal segment(s) of disease. More specifically, 25 patients (age: 60.67 ± 6.52 years, height: 1.77 ± 0.09 m, body mass: 90.50 ± 14.90 kg) had isolated AI disease. Four patients in the AI group had disease in one of their tibial arteries and the rest had no appreciable tibial disease. Twenty-three patients (age: 63.57 ± 6.69 years, height: 1.75 ± 0.08 m, body mass: 84.20 ± 19.66 kg) had isolated FP disease. Four patients in the FP group had disease in one, one had disease in two and two had disease in all three tibial arteries while the rest had no appreciable tibial disease. Forty-nine patients (age: 64.55 ± 6.11 years, height; 1.75 ± s0.05 m, body mass: 87.14 ± 17.17 kg) had MLD (combination of AI and FP disease). Eleven patients in the MLD group had disease in one, three had disease in two and one in all three tibial arteries while the rest had no appreciable tibial disease.

### Experimental Procedure and Data Collection

Prior to data collection, 27 retro-reflective markers were placed at specific anatomical locations on each participant’s lower limbs utilizing the marker systems of Vaughan and Nigg[37], [38]. To capture kinematic and kinetic gait parameters, each study participant was instructed to walk at their preferred walking speed over a 4.5-meter walkway containing in-ground force platforms. Participants with PAD performed overground walking trials in both a pain free and claudicating state. Each subject’s most affected leg based on lowest ABI value was used for analysis. During the pain free condition, a one-minute rest period was required between walking trials to ensure the absence of claudication; more rest was required if the participant indicated pain was present. To induce the claudicating state, each participant with PAD performed a six-minute walk test. For this test, participants were instructed to cover as much distance as possible in six minutes, walking around two cones placed 50 feet apart inside the laboratory. If the participant desired, they were able to rest during the test, but the six-minute timer continued to run. Confirmation that participants were experiencing claudication was obtained prior to performing the overground claudication trials. During the claudication condition, study participants were not given a one-minute rest between trials, and they confirmed the presence of claudication pain prior to beginning each trial.

### Data Analysis

#### Quality of life measures

Overall health and physical function were assessed using the WIQ[39] and SF-36[40]. The WIQ is a disease-specific questionnaire validated in patients with intermittent claudication. It consists of four subcategories: pain, walking distance, walking speed, and stair climbing ability. The SF-36 consists of eight health domains: physical function, limitation due to physical health, limitation due to emotional health, energy/vitality, general mental health, bodily pain, social function, and overall general health. The SF-36 has been extensively evaluated and tested with a variety of populations and is able to distinguish between groups of varying health status[41]. Both scales range from 0 to 100, with 100 being the higher functioning/more positive health outcome score.

#### Quantitative biomechanics measures

The participants’ lower extremity three-dimensional kinematics were collected using a Motion Analysis eight-camera motion capture system (Motion Analysis Corp, Santa Rosa, CA) sampling at 60Hz. Kinetic data were acquired using one Kistler piezoelectric force platform (Kistler North America, Amherst, NY) sampling at 600Hz. Motion capture data were filtered using a low pass Butterworth filter with a cutoff frequency of 5Hz. For the kinematic and kinetic data, a total of five clean foot contacts were collected and averaged for each study participant in each walking condition. A clean foot contact was defined as a step in which the foot in stance phase only came in contact with one force platform and no other contacts occurred on that platform during the walking trial. Spatiotemporal gait parameters were calculated by averaging data from five of the overground walking trials. Because spatiotemporal gait parameters were not initially analyzed for this data, two of the participants in the FP group and three healthy controls did not take enough steps for spatiotemporal characteristics to be calculated from each of the walking trials. Data were exported and processed using Cortex C-Motion Capture system (Motion Analysis Corporation, Santa Rosa, CA), Visual 3D (Germantown, MD, USA), and personalized MATLAB (The Mathworks, Inc., Natick, MA, USA) codes.

Joint kinematics and kinetics were calculated in the sagittal plane during the stance phase of walking. The stance phase can further be divided into three distinct stages: weight acceptance, single limb support, and push-off. An inverse dynamics solution was performed to calculate joint torques and powers from joint kinematics and ground reaction force data. Only data from each patient’s most affected leg were analyzed, and control participants were distributed to match the ratio of right to left legs that were analyzed from the patients with PAD. Torque and power data were normalized to each participant’s body mass.

A canonical linear discriminant function analysis (i.e., discriminant analysis) was performed to determine if differences in gait parameters could be used to distinguish between different levels of disease and healthy controls. Data were checked for multicollinearity to reduce the amount of mutual information in the model by running a bivariate Pearson’s correlation for both the pain and pain free data. During the pain free condition ankle plantarflexor torque, knee flexion torque, hip extension torque, ankle power absorption during weight acceptance, ankle power absorption during single limb support, ankle power generation during push-off, knee power generation during single limb support, hip power generation during weight acceptance, hip power absorption during single limb support, hip power generation during push-off, ankle range of motion, hip range of motion, stride length, and stance time were included in the analysis. During the pain condition, ankle dorsiflexion torque, knee flexion torque, hip extension torque, hip flexion torque, ankle power absorption during weight acceptance, ankle power absorption during single limb support, ankle power generation during push-off, hip power generation during weight acceptance, knee power absorption during push-off, ankle range of motion, knee range of motion, and stride time were included in the analysis.

### Statistical Analysis

Independent, one-way analysis of variance (ANOVA) tests were used to identify differences between participant groups (controls, AI, FP, MLD) for SF-36 subscale scores, WIQ subscale scores, and quantitative gait measures. A significant main effect indicates one or more significant differences between two or more of the four groups being compared. For variables with significant main effects, Bonferroni post hoc analyses were performed when necessary to determine which groups were significantly different from each other. Independent samples t-tests were used to compare controls to all PAD patients pooled in a single group. A discriminant analysis was performed for each walking condition to determine if gait impairments were able to distinguish between patients with different levels of disease and healthy controls and determine which gait variables were most useful in this calculation. All analyses were performed using SPSS 23 statistical software (SPSS Inc., Chicago, Ill) with an alpha level set to 0.05.

## RESULTS

### Demographics

Demographic and anthropometric data for each participating group can be found in Table I. There was not a significant between-groups effect of age, height, body mass, or body mass index. Ankle brachial index was significantly different between patient groups for both the most affected (p < 0.001) and least affected leg (p < 0.001). Patients with MLD had significantly lower ABI values in both the most and least affected legs compared to the AI (most affected: p < 0.001, least affected: p < 0.001) and FP (most affected: p = 0.019, least affected: p = 0.015) groups. Of the 25 patients with AI disease eight had mild, eleven minimal, and six no FP occlusive disease. Three of the patients with AI disease presented with calf claudication, five with thigh and calf, six with buttock and calf, nine with buttock, thigh, and calf, and two with buttock and thigh symptoms. Of the 23 patients with FP disease eleven had mild, nine minimal, and three no AI occlusive disease (two of these three patients had mildly ectatic AI segment). All 23 patients with FP disease presented with calf only claudication. All 49 MLD patients had combination of AI and FP disease. Fifteen presented with calf claudication, thirteen with thigh and calf, six with buttock and calf, fourteen with buttock, thigh, and calf, and one with buttock-only symptoms.

**Table I.** Demographic and anthropometric characteristics

| Clinical characteristics | Controls<br>(n = 35) | Aortoiliac<br>(n = 25) | Femoropopliteal<br>(n = 23) | Multi-level<br>(n = 49) |
| --- | --- | --- | --- | --- |
| Age (years) | 65.23 ± 9.51 | 60.67 ± 6.52 | 63.57 ± 6.69 | 64.55 ± 6.11 |
| Body mass (kg) | 79.90 ± 13.80 | 90.50 ± 14.90 | 84.20 ± 19.66 | 87.14 ± 17.17 |
| Height (m) | 1.74 ± 0.08 | 1.77 ± 0.09 | 1.75 ± 0.08 | 1.75 ± 0.05 |
| BMI | 26.37 ± 3.71 | 29.03 ± 5.59 | 27.37 ± 5.08 | 28.33 ± 5.59 |
| Sex | 29M | 25M | 22M 1F | 49M |
| ABI Most affected limb | > 0.90 | 0.63 ± 0.15<sup>b</sup> | 0.54 ± 0.21<sup>b</sup> | 0.41 ± 0.19 |
| ABI Least affected limb | > 0.90 | 0.85 ± 0.17<sup>b</sup> | 0.77 ± 0.19<sup>b</sup> | 0.62 ± 0.21 |
| 6MWD (m) |  | 315.5 ± 100.6 | 320.9 ± 78.4 | 284.88 ± 103.8 |

a = significantly different compared to aortoiliac group
b = significantly different compared to multi-level group
BMI = body mass index
ABI = ankle brachial index

### Questionnaire comparisons between healthy controls and PAD groups

When comparing WIQ subscale scores across participant groups, there were significant main effects for each of the four subscale categories (WIQ Pain: p < 0.001, WIQ Distance: p < 0.001, WIQ Speed: p < 0.001, WIQ Stairs: p < 0.001; Table II). Healthy control participants had significantly higher WIQ scores across all four subscales when independently compared to each patient group (Table II).

**Table II.** Walking Impairment Questionnaire (WIQ) subscale scores

| WIQ subscale scores | Controls<br>(n = 35) | Aortoiliac<br>(n = 25) | Femoropopliteal<br>(n = 23) | Multi-level<br>(n = 49) |
| --- | --- | --- | --- | --- |
| WIQ: Pain | 95.71 ± 8.54 | 44.79 ± 24.98<sup>a</sup> | 57.07 ± 17.61<sup>a</sup> | 48.72 ± 21.69<sup>a</sup> |
| WIQ: Distance | 92.20 ± 15.99 | 24.12 ± 29.84<sup>a</sup> | 28.29 ± 26.06<sup>a</sup> | 16.74 ± 17.97<sup>a</sup> |
| WIQ: Speed | 80.37 ± 18.98 | 36.59 ± 25.04<sup>a</sup> | 27.93 ± 22.21<sup>a</sup> | 27.91 ± 25.06<sup>a</sup> |
| WIQ: Stair | 92.14 ± 15.97 | 38.89 ± 25.35<sup>a</sup> | 40.22 ± 28.47<sup>a</sup> | 30.95 ± 24.17<sup>a</sup> |

a = significantly different compared to controls
b = significantly different compared to the femoropopliteal group
c = significantly different compared to the multi-level group

When comparing SF-36 subscale scores across participant groups, significant main effects were present for physical function (p < 0.001), limitations due to physical health (p < 0.001), limitations due to emotional health (p = 0.002), energy (p < 0.001), emotional health (p < 0.001), pain (p < 0.001), social function (p < 0.001) and general function (p < 0.001; Table III). Post-hoc tests revealed control participants had significantly higher SF-36 scores in independent comparisons to each level of disease group for all subscales except the emotional health subscale where the AI group did not differ from healthy controls (Table III). This indicates healthy controls had better functioning that all PAD groups for nearly all health domains assessed by the SF-36. In the emotional health subscale, the AI group scored higher than the MLD group (p = 0.004), indicating better emotional health for those with AI level disease. In the pain subscale the MLD group scored significantly higher than the FP group (p = 0.009), meaning the MLD group subjectively had less pain than the FP group.

**Table III.** Medical Outcomes Study Short Form 36 Health Survey (SF-36) subscale scores

| SF-36 subscale scores | Controls<br>(n = 35) | Aortoiliac<br>(n = 25) | Femoropopliteal<br>(n = 23) | Multi-level<br>(n = 49) |
| --- | --- | --- | --- | --- |
| Physical function | 88.67 ± 13.89 | 38.33 ± 16.40<sup>a</sup> | 43.47 ± 16.82<sup>a</sup> | 38.27 ± 17.75<sup>a</sup> |
| Limitations due to<br>physical health | 91.94 ± 26.13 | 28.13 ± 37.09<sup>a</sup> | 39.13 ± 33.56<sup>a</sup> | 23.98 ± 34.60<sup>a</sup> |
| Limitation due to<br>emotional health | 89.57 ± 26.70 | 61.11 ± 41.32<sup>a</sup> | 52.17 ± 39.98<sup>a</sup> | 60.54 ± 41.76<sup>a</sup> |
| Energy | 86.50 ± 17.39 | 49.90 ± 21.72<sup>a</sup> | 53.48 ± 22.00<sup>a</sup> | 44.44 ± 21.38<sup>a</sup> |
| Emotional Health | 77.79 ± 11.55 | 69.83 ± 15.85<sup>d</sup> | 63.22 ± 23.45<sup>a</sup> | 53.67 ± 21.01<sup>a,b</sup> |
| Pain | 82.35 ± 13.72 | 52.71 ± 23.97<sup>a</sup> | 49.17 ± 17.04<sup>a,d</sup> | 65.64 ± 22.64<sup>a,c</sup> |
| Social function | 94.83 ± 13.16 | 66.15 ± 27.46<sup>a</sup> | 65.98 ± 26.43<sup>a</sup> | 67.86 ± 24.34<sup>a</sup> |
| General health | 77.35 ± 17.37 | 52.71 ± 18.71<sup>a</sup> | 56.96 ± 23.05<sup>a</sup> | 53.27 ± 15.96<sup>a</sup> |

a = significantly different compared to controls
b = significantly different compared to aortoiliac group
c = significantly different compared to femoropopliteal group
d = significantly different compared to multi-level group

### Pain free walking: healthy controls versus PAD groups

There were significant main effects between participant groups for kinematic and kinetic gait parameters when walking in the pain-free condition for 12 of the 18 biomechanics measures (ankle dorsiflexor torque: p = 0.004, knee extensor torque: p = 0.006, hip extension torque: p < 0.001, hip flexor torque: p< 0.001, ankle power absorption during weight acceptance: p < 0.001, ankle power generation during push-off: p < 0.001, knee power absorption during weight acceptance: p = 0.013, knee power generation during single limb support: p = 0.002, knee power absorption during push-off: p < 0.001, hip power absorption during single limb support: p = 0.001, hip power generation during push-off: p < 0.001, hip range of motion: p = 0.003; Table IV). The six variables that did not have significant differences during pain free walking included: ankle plantarflexor torque during push-off, knee flexor torque during single limb support, ankle power absorption during single limb support, hip power generation during weight acceptance, ankle range of motion, and knee range of motion (Table IV). There were similar significant main effects for spatiotemporal gait parameters during pain free walking, with seven of the eight dependent variables demonstrating significant differences (step length: p < 0.001, stride length: p < 0.001, walking velocity: p < 0.001, stance time: p < 0.001, stride time: p = 0.002, step width: p = 0.006, cadence: p = 0.001) Only swing time did not have a significant main effect during pain free walking (Table V).

**Table IV.** Quantitative gait biomechanics during pain free walking

| **Gait Parameter** | **Controls**<br>**(n = 35)** | **Aortoiliac**<br>**(n = 25)** | **Femoropopliteal**<br>**(n = 23)** | **Multi-level**<br>**(n = 49)** | **All PAD**<br>**(n = 97)** |
| --- | --- | --- | --- | --- | --- |
| **ADT** | -0.32 ± 0.10 | -0.24 ± 0.08<sup>a</sup> | -0.26 ± 0.11 | -0.25 ± 0.10<sup>a</sup> | -0.25 ± 0.10<sup>a</sup> |
| **APT** | 1.38 ± 0.19 | 1.33 ± 0.18 | 1.31 ± 0.13 | 1.26 ± 0.22<sup>a</sup> | 1.29 ± 0.19<sup>a</sup> |
| **KET** | 0.69 ± 0.21 | 0.52 ± 0.30 | 0.52 ± 0.23 | 0.50 ± 0.24<sup>a</sup> | 0.51 ± 0.25<sup>a</sup> |
| **KFT** | -0.14 ± 0.15 | -0.21 ± 0.22 | -0.17 ± 0.16 | -0.21 ± 0.20 | -0.20 ± 0.20 |
| **HET** | 0.85 ± 0.29 | 0.69 ± 0.21<sup>a</sup> | 0.59 ± 0.23<sup>a</sup> | 0.64 ± 0.21<sup>a</sup> | 0.64 ± 0.22<sup>a</sup> |
| **HFT** | -1.05 ± 0.21 | -0.88 ± 0.24<sup>a</sup> | -0.81 ± 0.20<sup>a</sup> | -0.81 ± 0.23<sup>a</sup> | -0.82 ± 0.23<sup>a</sup> |
| **A1** | -0.67 ± 0.29 | -0.46 ± 0.17<sup>a</sup> | -0.53 ± 0.19 | -0.42 ± 0.25<sup>a</sup> | -0.46 ± 0.22<sup>a</sup> |
| **K1** | -0.91 ± 0.39 | -0.71 ± 0.54 | -0.62 ± 0.40 | -0.61 ± 0.40<sup>a</sup> | -0.64 ± 0.44<sup>a</sup> |
| **H1** | 0.41 ± 0.29 | 0.36 ± 0.29 | 0.36 ± 0.25 | 0.38 ± 0.29 | 0.37 ± 0.28 |
| **A2** | -0.83 ± 0.25 | -0.79 ± 0.27 | -0.78 ± 0.30 | -0.81 ± 0.25 | -0.80 ± 0.26 |
| **K2** | 0.45 ± 0.22 | 0.29 ± 0.23<sup>a</sup> | 0.29 ± 0.14<sup>a</sup> | 0.31 ± 0.18<sup>a</sup> | 0.30 ± 0.19<sup>a</sup> |
| **H2** | -0.84 ± 0.26 | -0.66 ± 0.32 | -0.62 ± 0.20<sup>a</sup> | -0.61 ± 0.28<sup>a</sup> | -0.62 ± 0.27<sup>a</sup> |
| **A3** | 2.70 ± 0.69 | 2.23 ± 0.64<sup>a</sup> | 1.90 ± 0.45<sup>a</sup> | 1.92 ± 0.64<sup>a</sup> | 2.00 ± 0.61<sup>a</sup> |
| **K3** | -1.19 ± 0.47 | -0.82 ± 0.23<sup>a</sup> | -0.79 ± 0.28<sup>a</sup> | -0.78 ± 0.37<sup>a</sup> | -0.79 ± 0.32<sup>a</sup> |
| **H3** | 0.99 ± 0.23 | 0.76 ± 0.15<sup>a</sup> | 0.69 ± 0.23<sup>a</sup> | 0.72 ± 0.23<sup>a</sup> | 0.72 ± 0.21<sup>a</sup> |
| **AROM** | 18.78 ± 3.66 | 19.12 ± 4.17 | 19.29 ± 3.34 | 20.05 ± 3.44 | 19.63 ± 3.60 |
| **KROM** | 12.21 ± 3.67 | 11.49 ± 4.53 | 11.13 ± 4.80 | 11.17 ± 5.01 | 11.25 ± 4.80 |
| **HROM** | 40.10 ± 3.27 | 36.63 ± 5.37<sup>a</sup> | 36.33 ± 4.73<sup>a</sup> | 36.51 ± 5.22<sup>a</sup> | 36.50 ± 5.10<sup>a</sup> |

a = significantly different compared to controls
PAD = peripheral artery disease ADT = ankle dorsiflexor torque, APT = ankle plantarflexor torque, KET = knee extensor torque, KFT = knee flexor torque, HET = hip extensor torque, HFT = hip flexor torque, A1 = ankle power absorption during weight acceptance, A2 = ankle power absorption during single leg support, A3 = ankle power generation during push off, K1 = knee power absorption during weight acceptance, K2 = knee power generation during single leg support, K3 = knee power absorption during push off, H1 = hip power generation during weight acceptance, H2 = Hip power absorption during single leg support, H3 = hip power generation during push off, AROM = ankle range of motion, KROM = knee range of motion, HROM = hip range of motion

**Table V.**
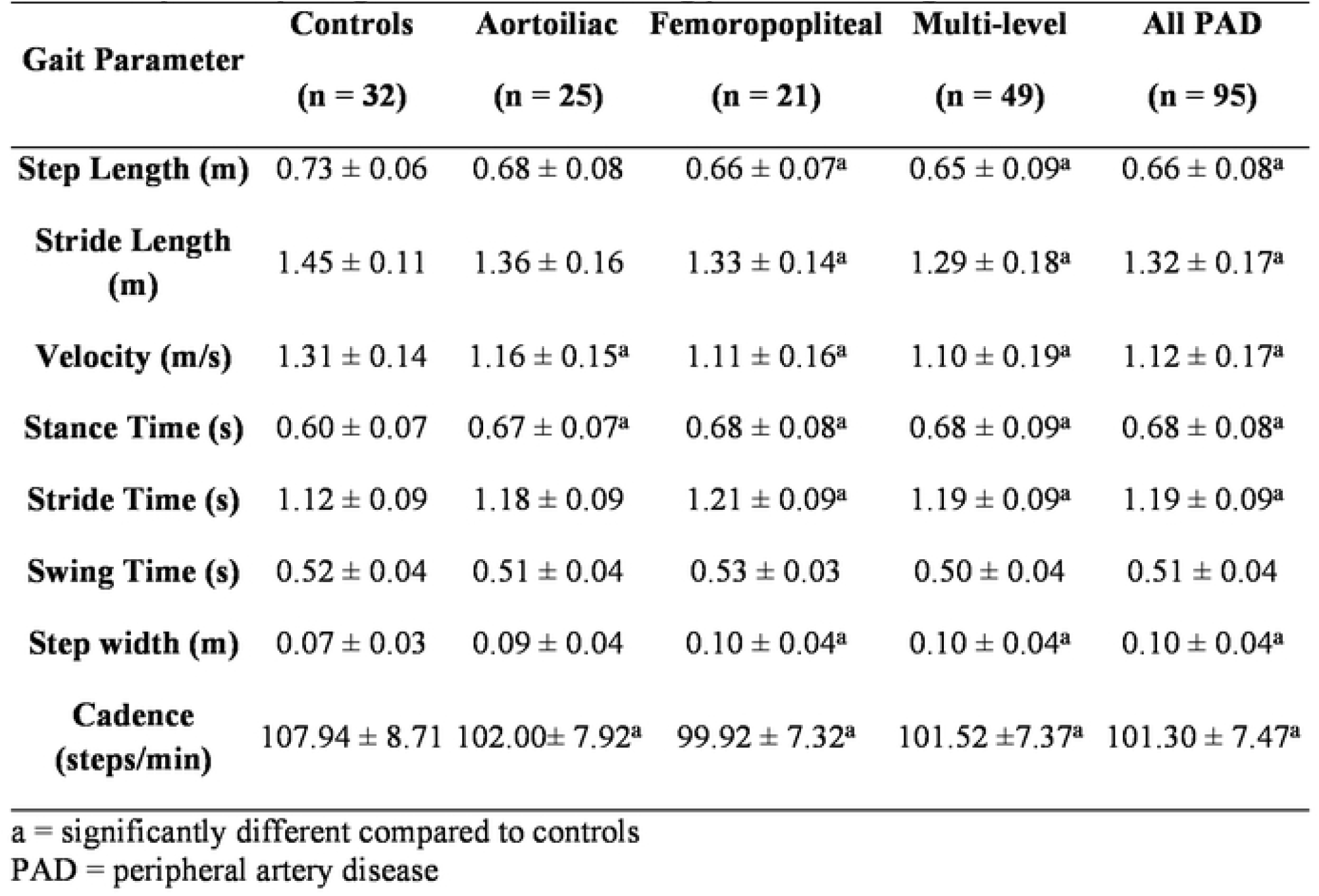
Spatiotemporal gait measurements during pain free walking

The post-hoc analysis revealed that all differences occurred between one or more level of disease groups and healthy controls. Bonferroni post hoc analyses indicated no differences for kinematic, kinetic, or spatiotemporal gait parameters between PAD level of disease groups when walking in the pain free condition. Of the total 26 gait comparisons (18 kinematic/kinetic and 8 spatiotemporal), the differences between healthy controls and all PAD groups occurred for 20 of 26 variables, patients with AI level disease for 12 of 26 variables, patients with FP for 15 of 26 variables, and patients with MLD for 20 of 26 variables (Tables IV and V).

### Pain walking: healthy controls versus PAD groups

When walking in the pain condition, there were significant main effects of participant group for 14 of 18 kinematic and kinetic gait parameters (ankle dorsiflexor torque: p < 0.001, ankle plantarflexor torque: p < 0.001, knee extensor torque: p = 0.008, hip extension torque: p < 0.001, hip flexor torque: p< 0.001, ankle power absorption during weight acceptance: p < 0.001, knee power absorption during weight acceptance: p = 0.007, knee power generation during single limb support: p = 0.005, hip power absorption during single limb support: p = 0.003, ankle power generation during push-off: p < 0.001, knee power absorption during push-off: p < 0.001, hip power generation during push-off: p < 0.001, ankle range of motion: p = 0.002, hip range of motion: p < 0.001). The four variables that did not show a significant main effect were knee flexor torque during single limb support, hip power generation during weight acceptance, ankle power absorption during single limb support, and knee range of motion (Table VI). There were also significant main effects for eight spatiotemporal gait parameters (step length: p < 0.001, stride length: p < 0.001, walking velocity: p < 0.001, stance time: p < 0.001, stride time: p = 0.006, step width: p = 0.009) Swing time and cadence did not show a significant main effect (Table VII).

**Table VI.** Quantitative gait biomechanics during claudication pain induced walking

| **Gait Parameter** | **Controls**<br>**(n = 35)** | **Aortoiliac**<br>**(n = 25)** | **Femoropopliteal**<br>**(n = 23)** | **Multi-level**<br>**(n = 49)** | **All PAD**<br>**(n = 97)** |
| --- | --- | --- | --- | --- | --- |
| **ADT** | -0.32 ± 0.10 | -0.22 ± 0.07<sup>a</sup> | -0.28 ± 0.11 | -0.22 ± 0.08<sup>a</sup> | -0.24 ± 0.09<sup>a</sup> |
| **APT** | 1.38 ± 0.19 | 1.29 ± 0.20 | 1.23 ± 0.14 | 1.16 ± 0.25<sup>a</sup> | 1.21 ± 0.22<sup>a</sup> |
| **KET** | 0.69 ± 0.21 | 0.56 ± 0.30 | 0.56 ± 0.25 | 0.50 ± 0.21<sup>a</sup> | 0.53 ± 0.24<sup>a</sup> |
| **KFT** | -0.14 ± 0.15 | -0.20 ± 0.23 | -0.17 ± 0.20 | -0.17 ± 0.19 | -0.18 ± 0.20 |
| **HET** | 0.85 ± 0.29 | 0.69 ± 0.20 | 0.58 ± 0.24<sup>a</sup> | 0.62 ± 0.22<sup>a</sup> | 0.63 ± 0.22<sup>a</sup> |
| **HFT** | -1.05 ± 0.21 | -0.88 ± 0.25 | -0.75 ± 0.25<sup>a</sup> | -0.73 ± 0.24<sup>a</sup> | -0.77 ± 0.25<sup>a</sup> |
| **A1** | -0.67 ± 0.29 | -0.46 ± 0.18<sup>a</sup> | -0.58 ± 0.30<sup>d</sup> | -0.36 ± 0.19<sup>a,c</sup> | -0.44 ± 0.23<sup>a</sup> |
| **K1** | -0.91 ± 0.39 | -0.76 ± 0.54 | -0.70 ± 0.46 | -0.58 ± 0.36<sup>a</sup> | -0.65 ± 0.44<sup>a</sup> |
| **H1** | 0.41 ± 0.29 | 0.38 ± 0.30 | 0.32 ± 0.25 | 0.34 ± 0.22 | 0.35 ± 0.25 |
| **A2** | -0.83 ± 0.25 | -0.81 ± 0.39 | -0.86 ± 0.38 | -0.86 ± 0.28 | -0.84 ± 0.33 |
| **K2** | 0.45 ± 0.22 | 0.32 ± 0.23 | 0.33 ± 0.21 | 0.29 ± 0.18<sup>a</sup> | 0.31 ± 0.20<sup>a</sup> |
| **H2** | -0.84 ± 0.26 | -0.72 ± 0.34 | -0.61 ± 0.27<sup>a</sup> | -0.63 ± 0.27<sup>a</sup> | -0.65 ± 0.29<sup>a</sup> |
| **A3** | 2.70 ± 0.69 | 2.07 ± 0.68<sup>a,d</sup> | 1.71 ± 0.55<sup>a</sup> | 1.60 ± 0.60<sup>a,b</sup> | 1.75 ± 0.64<sup>a</sup> |
| **K3** | -1.19 ± 0.47 | -0.83 ± 0.30<sup>a</sup> | -0.71 ± 0.31<sup>a</sup> | -0.69 ± 0.33<sup>a</sup> | -0.73 ± 0.32<sup>a</sup> |
| **H3** | 0.99 ± 0.23 | 0.78 ± 0.24<sup>a</sup> | 0.62 ± 0.27<sup>a</sup> | 0.63 ± 0.23<sup>a</sup> | 0.66 ± 0.25<sup>a</sup> |
| **AROM** | 18.78 ± 3.66 | 20.66 ± 4.82 | 21.45 ± 3.56 | 22.06 ± 3.49<sup>a</sup> | 21.56 ± 3.89<sup>a</sup> |
| **KROM** | 12.21 ± 3.67 | 12.94 ± 4.54 | 11.53 ± 5.63 | 10.78 ± 5.19 | 11.51 ± 5.17 |
| **HROM** | 40.10 ± 3.27 | 36.70 ± 5.37 | 36.11 ± 5.96<sup>a</sup> | 35.14 ± 5.23<sup>a</sup> | 35.77 ± 5.43<sup>a</sup> |

a = significantly different compared to controls
b = significantly different compared to aortoiliac
c = significantly different compared to femoropopliteal
d = significantly different compared to multi-level
PAD = peripheral artery disease ADT = ankle dorsiflexor torque, APT = ankle plantarflexor torque, KET = knee extensor torque, KFT = knee flexor torque, HET = hip extensor torque, HFT = hip flexor torque, A1 = ankle power absorption during weight acceptance, A2 = ankle power absorption during single leg support, A3 = ankle power generation during push, K1 = knee power absorption during weight acceptance, K2 = knee power generation during single leg support, K3 = knee power absorption during push off, H1 = hip power generation during weight acceptance, H2 = Hip power absorption during single leg support, H3 = hip power generation during push off

**Table VII.** Spatiotemporal gait measurements during claudication pain induced walking

| **Gait Parameter** | **Control**<br>**(n = 32)** | **Aortoiliac**<br>**(n = 25)** | **Femoropopliteal**<br>**(n = 21)** | **Multi-level**<br>**(n = 49)** | **All PAD**<br>**(n = 95)** |
| --- | --- | --- | --- | --- | --- |
| **Step Length (m)** | 0.73 ± 0.06 | 0.65 ± 0.09<sup>a</sup> | 0.65 ± 0.08<sup>a</sup> | 0.62 ± 0.09<sup>a</sup> | 0.63 ± 0.09<sup>a</sup> |
| **Stride Length**<br>**(m)** | 1.45 ± 0.11 | 1.30 ± 0.18<sup>a</sup> | 1.30 ± 0.17<sup>a</sup> | 1.22 ± 0.19<sup>a</sup> | 1.26 ± 0.18<sup>a</sup> |
| **Velocity (m/s)** | 1.31 ± 0.14 | 1.11 ± 0.19<sup>a</sup> | 1.08 ± 0.21<sup>a</sup> | 1.04 ± 0.20<sup>a</sup> | 1.07 ± 0.20<sup>a</sup> |
| **Stance Time (s)** | 0.60 ± 0.07 | 0.68 ± 0.08<sup>a</sup> | 0.71 ± 0.08<sup>a</sup> | 0.69 ± 0.10<sup>a</sup> | 0.69 ± 0.09<sup>a</sup> |
| **Stride Time (s)** | 1.12 ± 0.09 | 1.18 ± 0.10 | 1.22 ± 0.11<sup>a</sup> | 1.18 ± 0.13 | 1.19 ± 0.12<sup>a</sup> |
| **Swing Time (s)** | 0.52 ± 0.04 | 0.50 ± 0.04 | 0.52 ± 0.05 | 0.50 ± 0.04 | 0.51 ± 0.04 |
| **Step width (m)** | 0.07 ± 0.03 | 0.09 ± 0.04 | 0.10 ± 0.03<sup>a</sup> | 0.10 ± 0.04<sup>a</sup> | 0.10 ± 0.04<sup>a</sup> |
| **Cadence**<br>**(steps/min)** | 107.94 ± 8.71 | 102.00 ± 8.64 | 99.07 ± 8.76 | 103.14 ± 15.47 | 101.94 ± 12.66<sup>a</sup> |

a = significantly different compared to controls
PAD = peripheral artery disease

Bonferroni post hoc analyses indicated no differences for kinematic, kinetic, or spatiotemporal gait parameters between any PAD level of disease groups when walking in the pain condition except two differences. Those differences include the MLD group, which had a significantly larger ankle power absorption during weight acceptance compared to the FP group (p = 0.003), and the AI group produced significantly more ankle power during push-off compared to the MLD group (p = 0.019). All other differences occurred between one or more level of disease groups and healthy controls. The controls versus all PAD group comparison yielded differences in 21 of 26 gait variables. Controls were significantly different than patients with AI level disease for nine of 26 variables, patients with FP disease for 13 of 26 variables, and patients with MLD for 19 of 26 variables (Tables VI and VII).

### Classifying subject group from gait impairments

In the pain free condition, a total of 99 study participants were included in the discriminant analysis. Thirty-three subjects were removed from the total sample of 132 due to the inability to calculate spatiotemporal data or if they were considered to be outliers. Significant connections between the studied gait variables and subject grouping were found (p = 0.011, Wilk’s Lambda = 0.478). The classification results indicated the discriminant function correctly classified 56.6% of subjects into the correct subject group. The analysis was most accurate in predicting healthy controls with a 76.0% classification accuracy. This was followed by MLD, AI, and FP with a 65.7%, 36.8% and 35.0% classification accuracy respectively. The gait parameter with the most significant contribution to the prediction of subject group in the pain free condition was ankle power generation with a standardized canonical discriminant function coefficient of 0.504. The canonical correlation for this function was 0.629.

In the pain condition, a total of 105 study participants were included in the discriminant analysis. Similar to the pain-free conditions, 27 subjects were removed from the total 132 subjects due to the inability to calculate spatiotemporal data or if they were considered to be outliers. Significant connections between the studied gait variables and subject grouping were found (p < 0.001, Wilk’s Lambda = 0.424). The classification results indicated the discriminant function correctly classified 70.5% of subjects into the correct subject group. The analysis was most accurate in predicting the MLD group with an 88.6% classification accuracy. This was followed by controls, FP, and AI with a classification accuracy of 83.3%, 44.4% and 36.8% respectively. The gait parameter with the most significant contribution to the prediction of subject group in the pain condition was ankle power generation with a standardized canonical discriminant function coefficient of −0.539. The canonical correlation for this function was 0.658.

## DISCUSSION

Our data using advanced biomechanical analyses and standard clinical and quality of life assessments, indicate that different levels of arterial disease present with very different patterns of symptoms affecting the different muscle groups of the lower extremity but produce similar degrees of impairments across all muscle groups of the leg and in the walking patterns and walking distances of claudicating PAD patients. We found that PAD patients with different levels of vascular stenosis/occlusion have very similarly affected gait patterns (as described by kinematic, kinetic, and spatiotemporal gait parameters) and walking distances (measured with six-minute walk test) and very similar subjective impairments (measured with WIQ and SF-36). Patients with AI, FP and MLD disease all had similarly impaired biomechanics, compared to control, around their ankle, knee and hip joints (the muscle groups flexing and extending the ankle, knee and hip are similarly impaired in all PAD groups). None of the investigated biomechanical parameters differed between level of disease groups in the pain free walking condition, with minimal differences occurring in the claudicating condition. Moreover, when trying to determine if patients can be classified according to their level of disease based on gait impairments, and if quality of life information acquired through the WIQ and SF-36, differed across groups of patients with different levels of disease, we found that the model performed well in identifying healthy controls in both the pain (83.3% classification accuracy) and pain free (76.0% classification accuracy) conditions and that the model can also accurately classify individuals in the MLD group during the pain condition (88.6% classification accuracy), but had trouble differentiating between PAD subject groups in all other scenarios. Previous work has shown that gait impairments are present from the first steps a PAD patient takes while walking pain-free and are augmented when the patient starts experiencing claudication symptoms [10], [11]. This augmentation of walking impairments, especially in what is traditionally the lowest functioning group (MLD)[19], may help explain why the discriminant analysis was more accurate in identifying subject groupings during the pain condition compared to pain-free walking. The conclusion of no differences between level of disease groups is reinforced by the fact that all WIQ subscales scores were statistically similar between PAD groups. The SF-36 scores were also similar across level of disease groups with the only differences identified being in the subscales of pain between the FP and MLD groups and emotional health between the AI and MLD groups.

The results of the current study are not in accord with our hypothesis that different levels of occlusive disease uniquely affect the muscle contributions within the PAD legs and therefore produce distinctive differences in the way claudicating patients walk. Although it may be understandable that claudication produced by occlusions that are similar in hemodynamic significance impair walking distances and quality of life parameters in a similar fashion irrespective of the level of the arterial tree involved, it is difficult to explain how this would also be the case for gait biomechanics. For example, it is difficult to understand how a patient with FP disease presenting with calf claudication can have similarly affected knee and hip biomechanics reflecting similarly compromised thigh and buttock muscle contributions as a patient with AI or MLD disease presenting with calf, thigh and buttock claudication. However, this is exactly what our data demonstrate. More specifically all patients with FP disease present with claudication symptoms affecting only their calves while 22 out of 25 AI patients and 34 of 49 MLD patients present with claudication symptoms that also affect the thigh and/or buttock musculature, yet gait biomechanics were similar across the three groups. Our findings on level of symptoms confirms the well-known fact that different levels of arterial occlusive disease produce symptoms in different muscle groups (calves, thighs, buttocks) of the leg [17], [18] which led us to theorize that different levels of occlusive disease produce ischemia, myopathy and functional compromise of different muscle groups and this would result in distinctive biomechanical patterns around the ankle, knee and hip joints. The absence of difference in gait biomechanical patterns was surprising to us and raises several considerations that may explain our findings.

A possible explanation may be that AI, FP and MLD all affect the calf and foot of the patient and that the affected foot and calf dominate the biomechanical behavior of the entire leg [42]. This can lead to ankle movement impacting knee and hip mechanics. The stance phase of walking is a closed chain movement, so movement impairments at the ankle must produce a corresponding response (and possibly an impairment) at the knee and/or hip. Additionally, the biarticular nature of certain hamstring, quadricep, and calf muscles, makes it likely that pain in the calves, thigh, or buttocks can impact mechanics at more than one joint.

It is also possible that what we consider to be hemodynamically insignificant (<30% diameter reduction) disease in an arterial segment is a significant contributor to the functional deterioration of the muscles supplied by this arterial segment. This may be because the disease becomes hemodynamically significant (either pressures drop or flow patterns change across the segment) when exercise initiates higher flows through the diseased segment, or because the atherosclerotic plaques (even though they are mild or minimal) produce cytokines, reactive oxygen species and pro-inflammatory products that then reach and adversely affect the corresponding muscle groups [43], [44]. This possibility may be of particular interest since the vast majority of patients with FP disease have atherosclerotic changes in their AI segment (eleven had mild, nine minimal and only three had no occlusive disease) and similarly, the majority of AI patients have atherosclerotic changes in their FP segment (eight had mild, eleven minimal and six no femoropopliteal occlusive disease) even though these changes are not hemodynamically significant (<30%). Essentially, one of the most important findings of our study is the very low prevalence of strictly one-level of PAD in claudicating patients. Our data suggest that PAD is an atherosclerotic process that presents as multilevel disease with certain arterial segments becoming involved with more hemodynamically significant stenoses than others and this multilevel disease affects the ischemic legs of PAD patients in a multilevel fashion impairing the performance of all leg muscles at the buttock, thigh, and calf level.

It is also possible that occlusive disease in a segment of the arterial tree changes the flow patterns in the segment above it and that such disturbed flow patterns may affect the health of the endothelium and all the layers of the arterial wall of the seemingly “normal” arterial segment and the muscles that are perfused by the segment. Related to this is the recently described increase in arterial stiffness in the arteries of PAD patients[45]–[47]. This may be promoted by disturbed flow which stimulates pro-inflammatory responses from endothelial cells and promotes atherosclerotic remodeling of affected arteries through lipid accumulation and elastin degradation[46]. The work by Abraham et al. supports this possibility because it demonstrates a substantial decrease in the resting and post exercise transcutaneous oxygen saturation of the skin over the buttocks both in patients with AI disease but also in a large number of patients without AI disease[48], [49].

Finally, while it is important to focus on the downstream damage PAD produces to the tissues of the affected lower limbs (due to ischemia and ischemia/reperfusion of the leg) we should also consider the, thus far poorly explored, effects of PAD to the rest of the body (mediated through the activation of metabolic, neural, or inflammatory pathways) as this may also explain some of our findings. In this process the ischemic limb distributes and communicates to the rest of the body, the adverse ischemic events it suffers several times during the day, every time the patient experiences leg ischemia with or without associated claudication symptoms. In this process a patient with FP disease and an ischemic calf can experience pathological changes and decreased performance of the ipsilateral non-ischemic thigh or pelvic musculature but also damage to other limbs and organs of the body. This may be an important pathway explaining how PAD can produce the increased morbidity and mortality that is so well documented for PAD patients [5]– [7]. The best evidence for a systemic effect of PAD comes from work showing that a single bout of exercise of the PAD legs produces a significant increase of biomarkers of oxidative stress (increased malondialdehyde, consumption of anti-oxidants)[50], increased inflammatory cells (white blood cells and neutrophils)[51], [52] and increased inflammatory cytokines (thromboxane, p-selectin, von Willebrand factor) in the blood and a significant activation of the sympathetic system of PAD patients. Of note, the group of Sinoway and Cauffman have shown in several elegant studies that it takes a few, low-intensity, contractions of the posterior calf muscles of PAD patients to produce significant sympathetic system activation and adverse effects on physiologic parameters like the heart rate, blood pressure and coronary and renal artery blood flow[53]–[55]. It is possible that sympathetic system activation by ischemia of the calf can adversely affect the blood flow and the health of the thighs and buttocks. It is likely that the increased local production of anaerobic metabolites, reactive oxygen species, and cytokines is a key mediator for the activation of the sympathetic loops that originate in the ischemic leg [56], [57]. These loops convey the injurious information to the brain, the limb itself, and the rest of body including limbs and organs [58].

Ankle power generation at push-off was the most significant contributor in determining subject classification (control versus PAD including AI, FP, or MLD) in both the pain and pain free conditions. This is likely due to the importance of the ankle plantarflexors in propelling the individual forward during walking. The plantarflexor muscles contribute approximately 50% of the positive work on the body’s center of mass during propulsion and more mechanical power than the hip and knee joints combined[42], [59]–[61]. Similar to previous studies, our results indicate patients with PAD display significantly lower power and torque outputs at the ankle compared to healthy controls, with augmented differences after the onset of claudication[10], [11], [14], [15], [62]. However, still no differences between patient groups were observed for this variable. Although all PAD groups had vascular stenosis/occlusion at or above the knee joint, resulting in all groups experiencing inadequate blood flow to the plantarflexor muscles, research has shown oxygen delivery to the muscles is not the only limiting factor in exercise performance for individuals with PAD[63]. Our group and others have extensively described that atherosclerotic blockages produce a state of ischemia and ischemia/reperfusion in the PAD legs which initiates oxidative stress/damage, mitochondrial dysfunction and cytokine upregulation, producing injury to all tissues including muscles, nerves, skin, and subcutaneous tissues [20], [31], [64]. Accumulating injury in the leg leads to progressive damage of muscle structure and function which in association with exercise-induced ischemia produce the limitation in walking ability known as claudication. The work of several laboratories including our own has demonstrated that this myopathy is closely related to leg function, daily activity, quality of life and mortality of PAD patients [7], [65]–[68].

One possible limitation to this study is how level of disease was assigned to each participant. Participants were assigned to a disease group based on the level of the arterial tree which had significant stenosis/occlusion (>30% diameter reduction) as determined by vascular surgeons using computerized tomographic angiographic imaging[35] [36]. Many of these participants had atherosclerotic disease in other areas of the leg but this disease was minimal (less than 10% diameter reduction) or mild (less than 30% diameter reduction). However, given the study results, the authors do not believe this could have changed the results of the study given no differences were found between any of the PAD groups.

## CONCLUSION

Our data using advanced biomechanical analyses and standard clinical and quality of life assessments, indicate that different levels of arterial disease present with symptoms affecting different muscle groups of the lower extremity, but produce similar degrees of dysfunction in all muscle groups of the leg and in the walking patterns, distances, and impairments of PAD patients with claudication. PAD affects claudicating legs in a diffuse manner irrespective of the level of the arterial tree primarily involved. Several pathophysiologic parameters that have to do with the true nature of PAD as a multilevel disease and its complicated hemodynamics, neuropathy, myopathy, and systemic effects will need to be further researched to understand the diffuse manner in which PAD affects the legs of claudicating patients.

## Data Availability

All relevant data are within the manuscript and its supporting information files.

## Acknowledgements

This work was supported by grants from the National Institute of Health (R01AG034995, R01HD090333, R01AG049868) and the United States Department of Veterans Affairs Rehabilitation Research and Development Service (I01RX003266).

## Notes

### Competing Interest Statement

The authors have declared no competing interest.

### Funding Statement

IIP: R01 AG034995, R01AG049868, both National Institutes of Health, nih.gov, SAM: R01HD090333 National Institutes of Health, I01RX003266 US Department of Veterans Affairs Rehabilitation Research and Development Service. The funders had no role in study design, data collection and analysis, decision to publish, or preparation of the manuscript.

### Author Declarations

The Internal Review Boards of the University of Nebraska Medical Center and Omaha VA Medical Center reviewed and approved this research. Written informed consent was obtained from every subject involved in this work.

